# First Detection of Oropouche Virus in *Culicoides insignis* in the Ucayali Region, Peru: Evidence of a Possible New Vector

**DOI:** 10.1101/2024.12.06.24318268

**Authors:** Edwin Requena-Zúñiga, Miriam Palomino-Salcedo, María P. García-Mendoza, Maribel D. Figueroa-Romero, Nancy S. Merino-Sarmiento, Oscar Escalante-Maldonado, Ana L. Cornelio-Santos, Patricia Cárdenas-Garcia, Carlos Augusto Jiménez, César Cabezas-Sanchez

## Abstract

This report details the first detection of the Oropouche virus (OROV) in *Culicoides insignis* in the Ucayali region (Peruvian Amazon) during an outbreak study in 2024. Captures conducted in peri-urban and rural areas showed that *C. insignis* accounted for 96.7% of the collected *Culicoides* specimens. RT-qPCR testing confirmed the presence of the virus in two *C. insignis* pools, establishing this species as a potential OROV vector in the study area, thereby contributing to the understanding of the virus’s epidemiology in the Amazon region.

## Introduction

Arboviruses represent a significant public health issue in tropical and coastal areas of Peru, including diseases such as Dengue, Chikungunya, Mayaro, and Oropouche fever. The circulation of the Oropouche virus (OROV) is likely underestimated due to clinical similarities with Dengue fever and other arboviruses^(1)^.

Over the last 45 years, numerous OROV fever outbreaks have been reported across the Americas, posing a significant public health concern in tropical regions. The virus has been isolated in Brazil, Panama, Peru, and Trinidad and Tobago^(2)^. Between 2023 and 2024, 1,066 cases of Oropouche fever were reported in the Amazonas state of Brazil, with 699 cases in Manaus^(3)^. In Peru, 94 cases were recorded between 2016 and 2022 across six departments: Ayacucho, Cajamarca, Cusco, Loreto, Madre de Dios, and San Martín^(3)^. As of epidemiological week 8 of 2024, 146 cases had been registered in Loreto, Ucayali, and Madre de Dios^(4)^.

*Culicoides paraensis* is considered the primary urban and peri-urban vector of OROV. This small blood-feeding midge, measuring 1–2.5 mm, can be identified by specific wing pigmentation patterns useful for taxonomic purposes. These midges belong to the Diptera order, Nematocera suborder, and Ceratopogonidae family^(5)^.

*Culicoides insignis* is known for transmitting the Blue Tongue Virus (BTV), which affects sheep and wild ruminants^(6)^, and it could be a potential vector for other orbiviruses and arboviruses^(7)^, although found on Peruvian farms, it has not been implicated with OROV transmission. Other species, such as *Ochlerotatus serratus* and *Psorophora cingulata*, are also considered vectors of OROV in its sylvatic cycle, involving birds and mammals (sloths and primates) as hosts. Additionally, *Culex quinquefasciatus* has been identified as an OROV vector in Brazil^(8)^.

## Study Design and Methods

In February of this year, a study was conducted to identify potential vectors responsible for OROV transmission in outbreak areas of the Ucayali region, specifically in the districts/localities of Alexander Von Humboldt (Padre Abad Province), and Campo Verde, Nueva Requena, and Manantay (Coronel Portillo Province). This time of year is characterized by rain and warm weather, with an average temperature of 25 °C and relative humidity of 80%. Mosquito and midge collections were carried out in rural areas (new settlements) and peri-urban zones with farms and fields. Sampled habitats included peri-urban farms, chicken coops, and pig and mixed pig/sheep farms (Figure 1). Collection was done using CDC light traps set from 17:00 to 07:00 the next day. Specimens were transported to the Regional Reference Laboratory of Ucayali and immobilized at 8 °C for taxonomic identification. Identified *Culicoides* and *Culicidae* were grouped into species-specific pools and preserved at −80 °C, then transported to the Viral Metaxenic Laboratory of the National Institute of Health (INS) for OROV detection through RT-qPCR.

**Fig 1.**
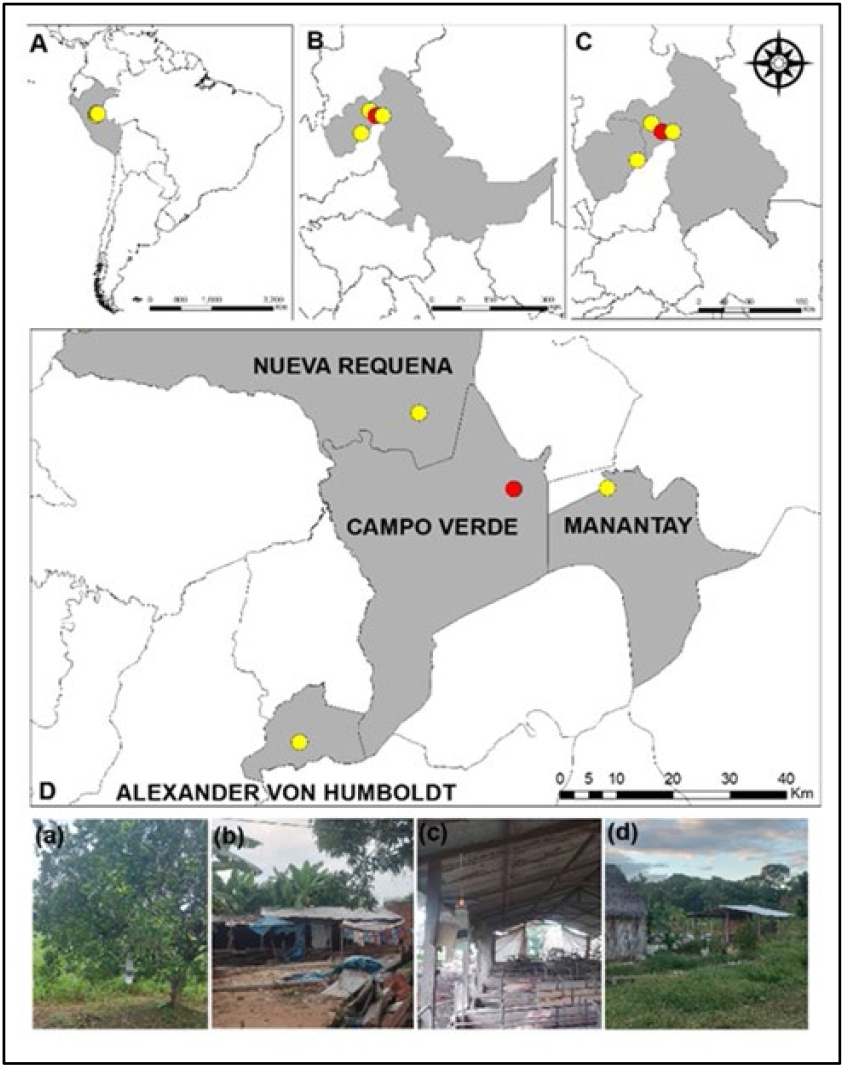
Map of the mosquito collection area in the Ucayali region, Peru. (A) South America; (B) Ucayali Region; (C) Padre Abad and Coronel Portillo Provinces; (D) Study area: The image shows the four sampled localities, with yellow dots representing the collection sites and the red dot indicating the positive OROV sample in Culicoides insignis; (a) Alexander Von Humboldt, (b) Manantay, (c) Campo Verde, (d) Nueva Requena.

## Results

A total of 2,583 mosquitoes and midges were collected, with 1,640 (63.5%) belonging to the *Culicoides* family, 96.7% of which were *C. insignis*, followed by 2.4% *C. leopoldoi* and 0.9% *C. foxi*. The remaining 36.5% were specimens from the *Anophelinae* and *Culicidae* families, spanning 10 genera (*Anopheles, Coquillettidia, Culex, Haemagogus, Mansonia, Ochlerotatus, Psorophora, Sabethes, Trichoprosopon*, and *Uranotaenia*).

## Virus Detection

Out of 2,567 identified female specimens, they were grouped into 117 pools (2 to 30 specimens each), with 56 pools for *Culicoides* (*C. insignis* and *C. leopoldoi*) and 61 pools for *Culicidae* species. *C. insignis* constituted 55 pools. RT-qPCR testing detected OROV positivity in two pools of *C. insignis* from Campo Verde.

## Conclusion

The *Culicoides* family, particularly *C. insignis*, can be considered a primary vector for OROV. This study’s detection of OROV in *C. insignis* suggests it as a potential vector in the peripheries of Pucallpa in the Ucayali region. Furthermore, *Culicoides* are recognized vectors of certain arboviruses causing severe diseases in domestic animals, such as Blue Tongue and African Horse Sickness. Continued vector incrimination and surveillance studies are necessary to guidecontrol measures in light of OROV’s spread in the Americas.

## Data Availability

not yet

## Acknowledgments

Thanks to Drs. Leticia Franco and Ana Cecilia Ribeiro of the PAHO/WHO Entomovirological Network and to Jhennyfert Custodio Mancilla, to Biologists Octavio Monteverde Daza and Layné Guerra Vargas (DIRESA Ucayali) for their technical support.

